# To test or not? Xpert MTB/RIF as an alternative to smear microscopy to guide line probe assay testing for drug-resistant tuberculosis

**DOI:** 10.1101/2022.12.05.22283088

**Authors:** S Pillay, M de Vos, H Sohn, Y Ghebrekristos, T Dolby, RM Warren, G Theron

**Author notes:** **Corresponding author:** Grant Theron, **Email:**, **Contact number**, **Address:** BMRI Building, 2^nd^ floor, Room 2035, Department of Molecular Biology and Human Genetics, Francie van Zijl Drive, Tygerberg Campus, Stellenbosch University, 7505, South Africa. Margaretha de Vos.

## Abstract

**Background:** Xpert MTB/RIF (Xpert) revolutionised tuberculosis (TB) diagnosis, however, laboratory decision making on whether widely-used reflex drug susceptibility assays (MTBDR*plus*, MTBDR*sl*) are done on specimens is often based on smear microscopy status.

**Method:** We performed receiver operator characteristic (ROC) curve analyses using sputum bacterial load measures [smear microscopy grade, Xpert semi-quantitation category and minimum cycle threshold (C_Tmin_) values] for the classification of “likely non-actionable” (not resistant or susceptible) line probe assays results. We evaluated the actionable-to-non-actionable result ratio and pay-offs with missed isoniazid and fluoroquinolone resistance compared to if LPAs were done universally.

**Findings:** Smear-negatives were more likely than smear-positives to generate a non-actionable MTBDR*plus* [23% (133/559) vs. 4% (15/381)] or MTBDR*sl* [39% (220/559) vs. 12% (47/381)] result, however, excluding smear-negatives would result in missed rapid diagnoses [e.g., only 51% (273/537) of LPA-diagnosable isoniazid resistance detected if smear-negatives omitted]. Within smear-negatives, testing ≥ “medium” specimens had a high ratio of actionable-to-non-actionable results (12.8 or a 4-fold improvement vs. test all for MTBDR*plus*, 4.5 or 3-fold improvement for MTBDR*sl*), which would capture 64% (168/264) and 77% (34/44) of LPA-detectable resistance. If C_Tmin_ were used, greater resolution and higher ratios offset against fewer missed resistant cases were obtained.

**Conclusion:** Routinely-generated Xpert quantitative information permits identification of smear-negatives in whom the ratio of actionable-to-non-actionable LPA results may prove acceptably high to laboratories depending on their local contexts. Xpert C_Tmin_ or, if unavailable, semiquantitation category should be used to guide reflex DST; permitting the rational expansion of direct DST to certain paucibacillary specimens.

## Introduction

Reflex drug susceptibility testing (DST) should be done in all rifampicin-resistant tuberculosis (TB) cases to enable rapid effective treatment. Achieving this depends on testing specimens, including smear-negative specimens, directly. However, the widely-used World Health Organization (WHO)-endorsed line probe assays (LPAs) MTBDR*plus* and especially MTBDR*sl* (both Bruker, Germany) perform sub-optimally on paucibacillary specimens (1) and can fail to generate an actionable (resistance or susceptible) result. Culture is hence often required to generate material for testing; however, culture is costly and slow.

Non-actionable MTBDR*plus* and MTBDR*sl* results in our setting occur in ∼24 and ∼40% of Xpert MTB/RIF (Xpert)-positive smear-negative patients, and are a bigger cause of missed resistance than diminished LPA sensitivity (2). Laboratories hence typically use smear status to guide whether LPA testing is done directly or indirectly and may choose not to test smear-negative specimens, however, this reduces rapid diagnoses. Alternatively, if LPAs are done on smear-negative specimens, wasteful expenditure (consumables and labour to do a MTBDR*sl* are ∼$50 (3)), care cascade loss (requests for additional specimen is typically only triggered once the LPA is known to be non-actionable are often unsuccessfully fulfilled), and reduced user confidence can all result if a non-actionable result occurs. Therefore, despite the WHO recommendation that MTBDR*sl* is done on smear-negative specimens, direct testing is often in reality limited to smear-positive specimens, even in well-resourced settings (4, 5). This undermines LPAs’ potential impact, which remain the only widely-deployed molecular DSTs for first- and second-line resistance. LPAs may indeed work well on some smear-negatives; however, as smear microscopy is a crude and insensitive categorical measure of bacterial load, laboratories are unable to identify this subset upfront prior to LPAs (6).

We hypothesised that, in situations where Xpert is a frontline TB test, its molecular quantitative information could be used to exclude *a priori* certain specimens from unnecessary LPA testing; thereby permitting LPAs to be applied more efficiently (i.e., on specimens with a reduced non-actionable result risk) and, if laboratories do not test smear-negative specimens, LPAs could be expanded to include some smear-negatives. In other words, pre-existing quantitative information routinely generated by Xpert could be used to improve LPA-based laboratory decision making and the drug-resistant TB care cascade. We also evaluate if smear grade would be more useful than smear status (positive, negative) for situations where Xpert is not available.

## Methods

### Microbiology

We analysed Auramine smear microscopy, Xpert MTB/RIF (v4.3), MTBDR*plus* and MTBDR*sl* (both v2) results from 951 patients programmatically-diagnosed with Xpert rifampicin-resistant TB from 01/06/2016-30/09/2019 at a high-volume laboratory in a previously-described cohort (2). All patients had sputum tested directly with both LPAs irrespective of smear status.

### Analyses

We did receiver operator characteristic (ROC) curve analyses (GraphPad v6, USA) using different sputum bacterial load measures to classify if MTBDR*plus* or MTBDR*sl* were non-actionable (not resistant or susceptible; defined as when bands corresponding to the amplification control or TB detection are absent or, if both present, ≥1 drug class locus control band was absent). Smear microscopy grade (per (7)), Xpert semi-quantitation category and minimum cycle threshold (C_Tmin_) values (rounded to nearest integer) were analysed, and sensitivity and specificity (95% binomial confidence intervals) for non-actionable results evaluated. We identified thresholds corresponding to Youden’s index (8), rule-out (≥95% sensitivity; almost all non-actionables correctly identified) and rule-in (≥95% specificity; almost all actionables correctly identified) scenarios; expecting rule-in to be most appropriate because it would not incorrectly exclude patients from the benefits of rapid LPA testing. We calculated, at each threshold, how many actionable results are generated before a non-actionable is encountered (ratio of actionable-to-non-actionable results) and how maximising this ratio was offset against missed LPA-based isoniazid and fluoroquinolone diagnoses.

### Ethics

This study was approved by the Health Research Ethics Committee of Stellenbosch University (N16/04/045) and Western Province Department of Health (2016/RP18/637).

## Results

### Non-actionable LPAs and missed resistance diagnoses stratified by smear status and grade

Non-actionable result rates irrespective of smear status for MTBDR*plus* and MTBDR*sl* were 19% (148/792) and 40% (267/673) (actionable-to-non-actionable results ratios of 5.4 and 2.5, respectively). Smear-negative specimens were, compared to smear-positives, more likely to generate a non-actionable MTBDR*plus* [23% (133/559) vs. 4% (15/381); p=0.001] or MTBDR*sl* [39% (220/559) vs. 12% (47/381); p<0.001] result (ratios of 3.2 and 24.4 for MTBDR*plus*, 1.5 and 7.1 for MTBDR*sl*, respectively). Non-actionable results, a receiver operating characteristic (ROC) curve of smear grade to detect non-actionable results, and the balance between the number of actionable results per non-actionable result and missed rapid drug resistance diagnoses are in **Figure 1** (positive and negative predictive values in **Supplementary Figure 1**).

**Figure 1.**
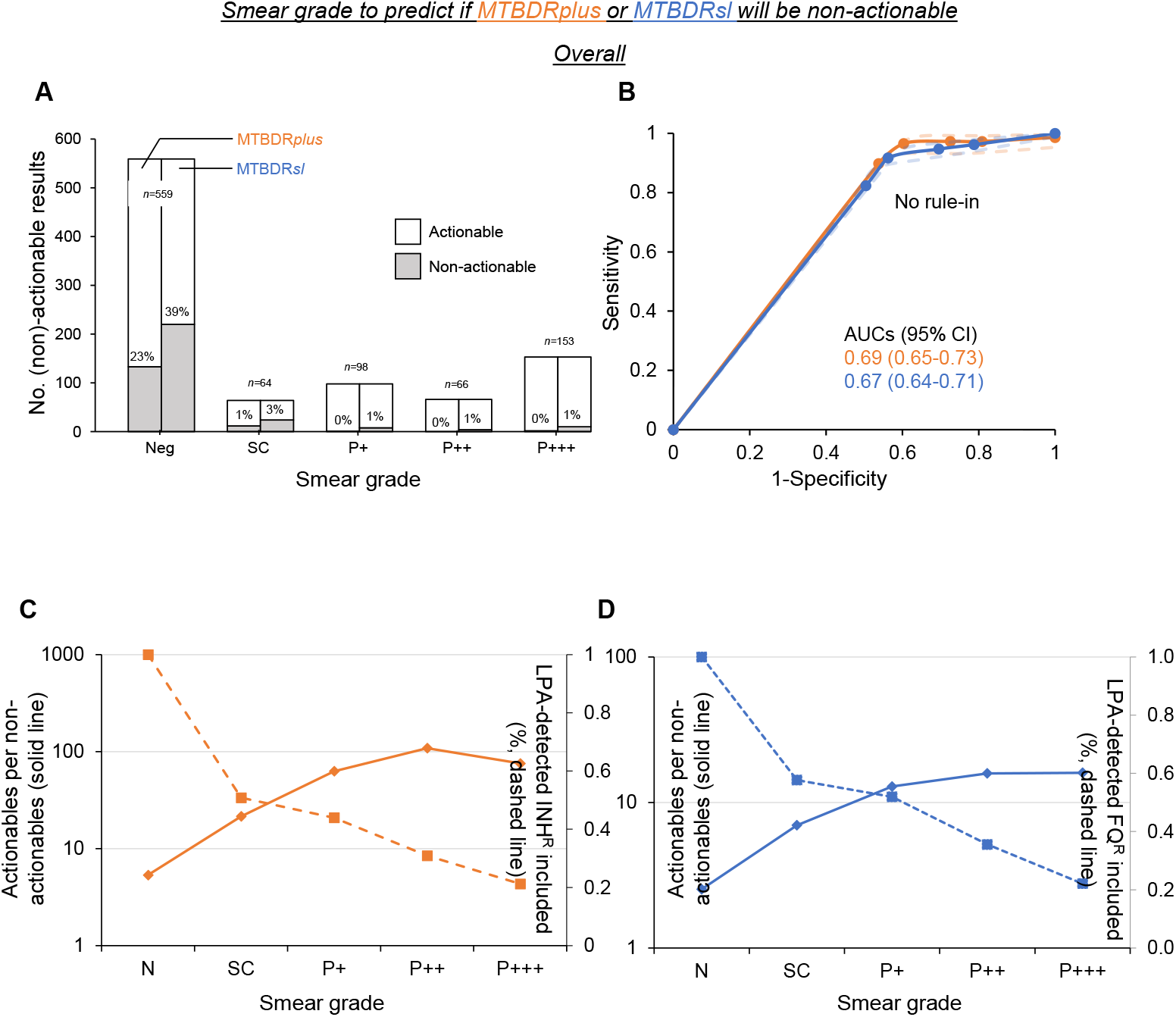
Smear grade’s association with non-actionable LPA results, its ability to discriminate “likely non-actionable” from “likely actionable” results (if ≤ each grade) and pay-offs between the ratio of actionable-to-non-actionable results with the overall proportion of LPA-detected resistance. **(A)** Non-actionable results were more frequent at lower than higher grades and more so for MTBDR*sl* than MTBDR*plus*. In-column percentages reflect the proportion patients with a non-actionable result. **(B)** Smear grade had moderate AUCs for identifying “likely non-actionable” results (dashed lines 95% CIs) but no grade approached 95% specificity. **(C)** and **(D)** show the ratio of actionable-to-non-actionable results (solid lines, left y-axes) and how this improves as specimens with a certain smear grade (or greater) are tested by MTBDR*plus* or MTBDR*sl* respectively. Abbreviations: AUCs-area under curve, CI-confidence intervals, FQ^R^-fluroquinolone resistance, INH^R^-isoniazid resistance, LPA-line probe assay, P-positive, SC-scanty, Xpert-Xpert MTB/RIF.

#### MTBDRplus

Smear-negativity as a threshold to identify non-actionables had a sensitivity and specificity of 90% (133/148) and 54% (426/792), respectively. Most non-actionable results occurred in smear-negatives (**Figure 1A**), but smear grade had suboptimal area under the curve (AUC) for predicting non-actionable results (**Figure 1B**). The actionable-to-non-actionable ratio improves as increasing grades are used to exclude specimens (≤that grade) from testing, however, this is offset against missed resistance (**Figure 1C**). For example, to improve this ratio to 21.5 (threshold ≤scanty or, in other words, any smear-positive tested), 51% (273/537) of LPA-diagnosable isoniazid resistance would be detected (**Supplementary Table 1**).

#### MTBDRsl

Smear-negativity had a sensitivity and specificity of 83% (220/266) and 54% (339/674) for non-actionable results. The actionable-to-non-actionable ratio was less than MTBDR*plus*’s, driven by more frequent non-actionable results in smear-negatives [39% (220/559) vs. 23% (133/559) for MTBDR*plus*, p<0.001]. For example, MTBDR*sl*’s highest ratio was 16 (**Figure 1D**) whereas for MTBDR*plus* it was 109 (∼7-fold higher). If smear-negative specimens were excluded from MTBDR*sl*, only 58% (60/104) of LPA-diagnosable fluroquinolone resistance would be detected (**Supplementary Table 1**).

### Xpert MTB/RIF semi-quantitation category

#### All patients

MTBDR*plus*: Like smear grade, non-actionable results were more frequent at lower semi-quantitation categories (**Figure 2A**), however, non-actionable rates in the “very low” and “low” categories were higher than in smear-negatives overall [49% (62/126) and 29% (62/210) in each category respectively vs. 23% (133/559; p<0.001 and p=0.103)]. Semi-quantitation category had higher AUC than smear grade, yet no semi-quantitation category threshold approached the rule-in criterion (∼95% specificity, **Figure 2B**). The largest improvement in the ratio of actionables-to-non-actionables occurred when specimens in the lowest two semi-quantitation categories were excluded (5.4 when all tested to 24.2 if ≥medium tested) and this was accompanied by a moderate reduction in detected resistance (22%) (**Figure 2C**). In other words, if ≥medium was used, ∼5-fold fewer non-actionables would occur and 78% (419/537) of potentially detectable resistance would still be detected.

**Figure 2.**
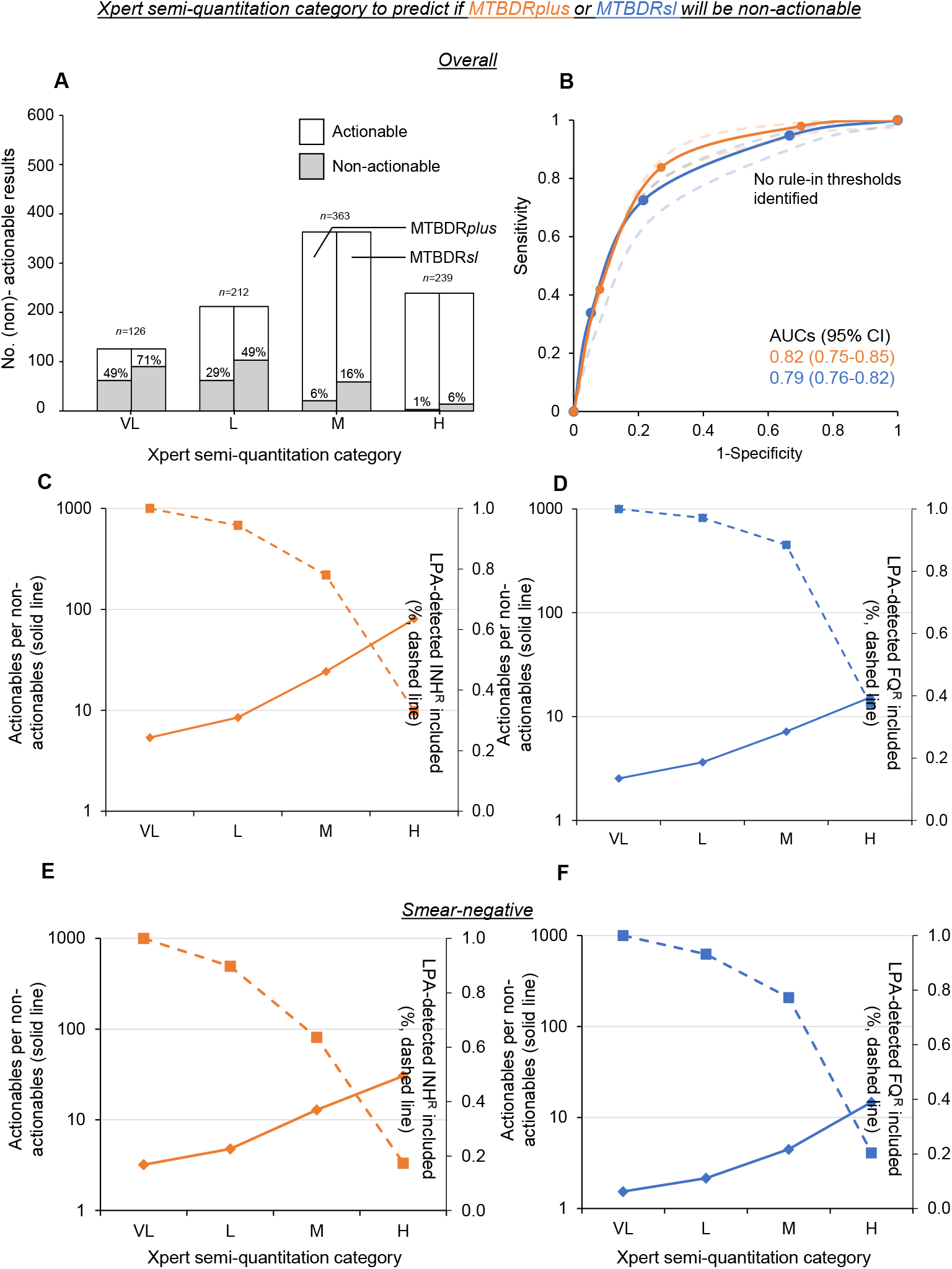
Xpert semi-quantitation category, non-actionable LPA results, and associated pay-offs with missed resistance as specimens ≤ specific semi-quantitation categories are excluded due to being flagged as “likely non-actionable”. **(A)** Trends for semi-quantitation category mirrored those for smear grade. **(B)** This translated into excellent AUCs for discriminating “likely non-actionable results” (dashed lines 95% CIs) but no optimal rule-in threshold was identifiable. **(C)** and **(D)** shows the ratio of actionable-to-non-actionable results (solid lines, left y-axes) and how this improves as specimens with higher semi-quantitation categories are tested by MTBDR*plus* or MTBDR*sl*, respectively. **(E)** and **(F)** are limited to smear-negative specimens. Abbreviations: AUCs-area under curve, CI-confidence intervals, FQ^R^-fluoroquinolone resistance, H-high, INH^R^-isoniazid resistance, LPA-line probe assay, L-low, M-medium, NPV-negative predictive value, P-positive, PPV-positive predictive value, VL-very low, Xpert-Xpert MTB/RIF.

MTBDR*sl*: Like MTBDR*plus*, MTBDR*sl* non-actionable rates in “very low” and “low” were higher than in smear-negative patients [71% (90/126) and 49% (102/210) in each category respectively vs. 39% (220/559; p<0.001 and p=0.021)]. MTBDR*sl* never obtained similar actionable-to-non-actionable ratios to MTBDR*plus* when specimens with the same semi-quantitation category were compared. Importantly, if ≥medium was used (specimens less than this excluded as “likely non-actionable), this ratio improved ∼3-fold from 2.5 to 7.2 with 88% (92/104) of potentially detectable resistance detected (**Figure 2D**).

#### Smear-negatives

MTBDR*plus*: If laboratories that do not test smear-negative patients wish to partly expand testing, they may test smear-negatives who are ≥medium (ratio 12.8 vs. 3.2 for the test all strategy or 4-fold improvement), which would still capture 64% (168/264) of detectable resistance (**Figure 2E)**. Within smear-negatives, 20% (114/559), 33% (182/559), 36% (200/559), and 11% (63/559) were “very low”, “low”, “medium”, and “high”, respectively; meaning 47% (263/559) of smear-negatives would be ≥medium (this rule could hence significantly expand testing in smear-negatives).

MTBDR*sl*: Similarly, if MTBDR*sl* was done on ≥medium smear-negatives, the ratio would improve from 1.5 for the test all strategy to 4.5 (3-fold improvement), with 77% (34/44) of detectable resistance (**Figure 2F)**.

### By Xpert MTB/RIF C_Tmin_

#### All patients

MTBDR*plus*: C_Tmin_ had, compared to Xpert semi-quantitation category and smear grade, higher AUC for non-actionable results **(Figure 3A)** and was the only bacillary load readout that met the rule-in criterion. 11% (86/792) of patients were C_Tmin_ ≥29. This threshold had 95% (750/792) specificity, meaning 5% (42/792) of actionables would be misclassified as “likely non-actionable” and hence excluded from MTBDR*plus* **(Supplementary Table 1)**. C_Tmin_ ≥29 sensitivity was 30% (44/148), meaning 44 non-actionables would be correctly classified as “likely non-actionable” (non-actionables reduced by a third). NPV was 80% (750/854) meaning that, for every ten patients C_Tmin_ <29 (hence classified as “likely actionable”), eight would indeed be actionable and other two non-actionable (false-negative). PPV was 51% (44/86), meaning approximately half of patients C_Tmin_ ≥29 (hence classified as non-actionable), would indeed be non-actionable and the others actionable (false-positive) **(Supplementary Figure 1)**. Ratios of actionable-to-non-actionable results like those for semiquantiation category were obtained, peaking at ∼77 (C_Tmin_ 12; estimates less than this were imprecise due to few specimens with very low C_Tmin_s) **(Figure 3B)**. At C_Tmin_ ≥29, this ratio would be 6.4 (improved from the test-all ratio of 5.4) and this would come at the cost of missing 5% (25/537) of potentially-detectable resistance.

**Figure 3.**
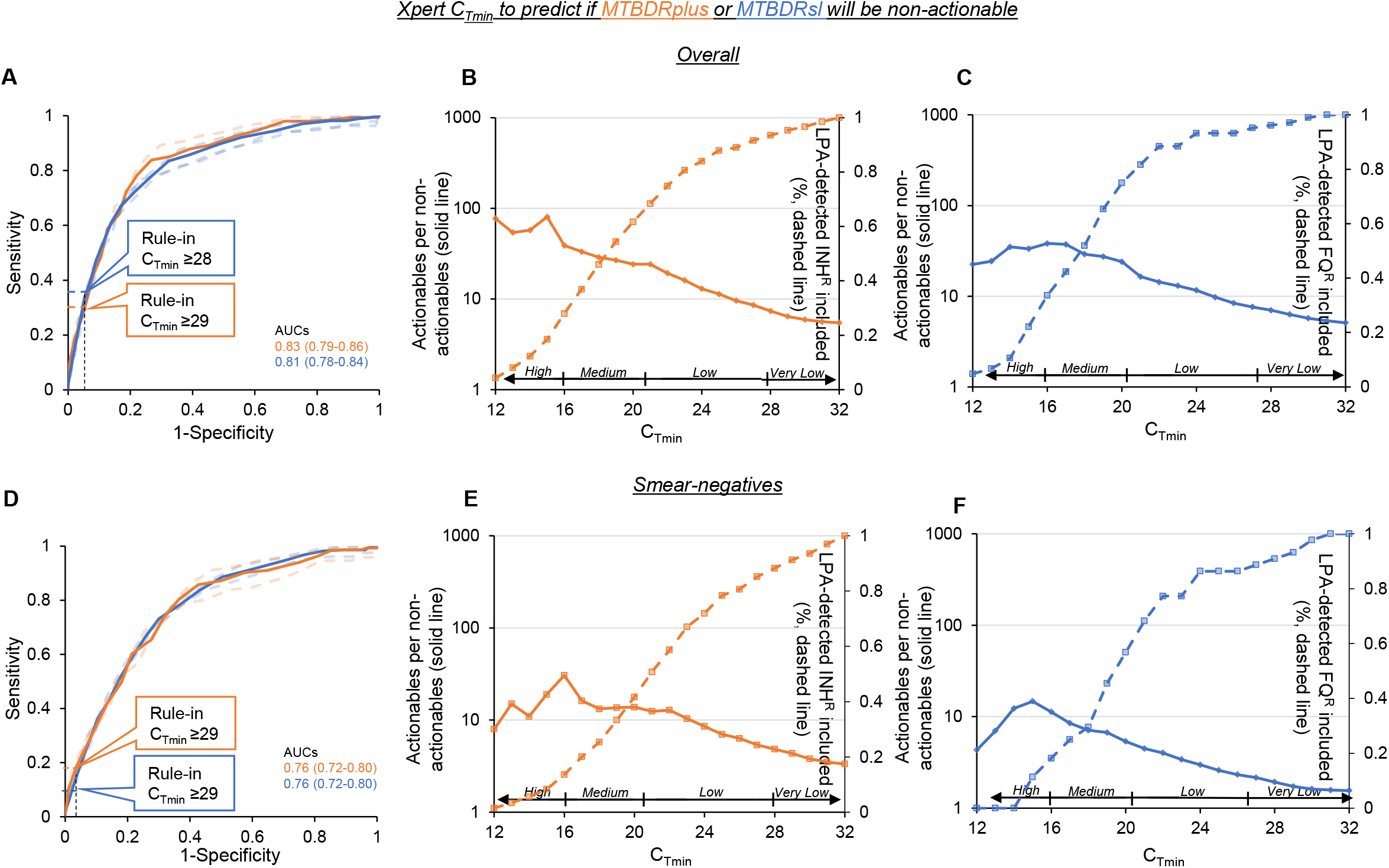
Xpert C_Tmin_’s ability to discriminate “likely non-actionable” from “likely actionable” LPA results. **(A)** A ROC curve for all specimens showing AUCs (dashed lines 95% CIs, rule-in thresholds shown) and, in **(B)** and **(C)**, pay-offs between the ratios of actionable-to-non-actionable results and missed resistance for MTBDR*plus* and MTBDR*sl*. **(D-E)** are the same but restricted to smear-negative patients. Ratios were highest at low C_Tmin_ and slowly decreased as LPA testing was expanded to include samples with higher C_Tmin_s, which had the upside of increasing detected resistance. AUCs and these ratios were less for smear-negative vs. all patients. Above C_Tmin_ x-axes are Xpert semiquantitation categories. Abbreviations: AUC-area under curve, CI-confidence intervals, C_Tmin_-cycle threshold (minimum), FQs-fluoroquinolones, INH-isoniazid, LPA-line probe assay, NPV-negative predictive value, P-positive, PPV-positive predictive value, ROC-receiver operator characteristic, Xpert-Xpert MTB/RIF.

MTBDR*sl:* 19% (129/674) of patients had C_Tmin_ ≥28, which had a rule-in specificity of 95% (638/674), meaning 5% (36/674) of actionables would be misclassified as “likely non-actionable” **(Figure 3A)**. Sensitivity was 34% (90/266); hence 90 non-actionables would be correctly classified as “likely non-actionable”, permitting a one third reduction in non-actionables. NPV was like that for MTBDR*plus* **(Supplementary Figure 1)** but PPV higher [71% (90/126; p=0.003 vs. MTBDR*plus*), meaning approximately 7/10 people with C_Tmin_ ≥28 (hence classified as non-actionable), would indeed be non-actionable and the other 3/10 actionable (false-positive). Ratios of actionable-to-non-actionables results peaked at ∼38 (C_Tmin_ 16), less than half that of MTBDR*plus*. At the C_Tmin_ ≥28 threshold, this ratio would be 7.0 (compared to the test-all ratio of 5.0, 1.4-fold or 40% improvement) and would result only 4% (4/104) of potentially detectable resistance being missed.

#### Smear-negative patients

MTBDR*plus*: Compared to overall, C_Tmin_ had less AUC in smear-negatives but similar rule-in threshold **(Figure 3D)**. Even at the same C_Tmin_s, lower actionable-to-non-actionable ratios occurred in smear-negatives **(Figure 3E**; for example, 13.8 vs. 24 overall at C_Tmin_ 20,). If the rule-in threshold of C_Tmin_ <29 was used, this ratio was 4.4 (compared to 3.2 for the test-all smear-negatives strategy, representing a 38% improvement) and resulted in 91% (241/264) of potentially detectable resistance captured. Furthermore, ratios ≥10 were possible, permitting MTBDR*plus* to be expanded to at least some smear-negatives (C_Tmin_ <23; 67% (177/264) of smear-negatives and 67% (177/264) of LPA-detectable resistance was C_Tmin_ <23.

MTBDR*sl*: If the rule-in threshold of C_Tmin_ <29 was used, this ratio was 1.7 (compared to 1.5 for the test-all smear-negatives strategy, a 13% improvement) and resulted in 93% (41/44) of potentially detectable resistance detected. MTBDR*sl* on specimens with C_Tmin_ <19 would have a ratio of 5.4 **(Figure 3F**), which may be more acceptable in settings where smear-negative testing is not routinely done. This ratio was more than the test-all strategy (3.6-fold improvement) and use of ≥medium semi-quantitation category (ratio of 4.5). 36% (119/332) of smear-negatives were C_Tmin_ <19, corresponding to 45% (24/44) of detectable resistance. Predictive values of this approach in smear-negatives, including for MTBDR*plus*, are in **Supplementary Figure 1**.

## Discussion

LPAs are WHO-recommended first- and second-line rapid DSTs, however, they are not always done directly on specimens in which they may provide an actionable resistant or susceptible result, in part due to elevated non-actionable result risk in smear-negatives. This can deprive patients of the benefits of early DST possible using presently available tests. Although better DSTs, especially for second-line resistance, are doubtlessly required, the use of existing widely-available technologies should be optimised.

Our key findings regarding smear-negative specimens are: 1) testing specimens below certain C_Tmin_ thresholds with MTBDR*plus* reduces non-actionable result rates and allows most LPA-detectable isoniazid resistance to be detected, 2) for MTBDR*sl*, which usually results in 1.5 actionables per non-actionable, ratios close to five are attainable (C_Tmin_ <19), permitting 45% of detectable fluroquinolone resistance to be detected, and 3) in settings where C_Tmin_s are unavailable, Xpert semi-quantitation category ≥medium would expand LPA testing to almost half of smear-negatives. Our study provides a framework for how LPA testing on smear-negatives can be made more efficient The precise threshold (and type of readout) used to determine whether LPA testing on smear-negatives should proceed will depend on locally-acceptable ratios of actionable-to-non-actionable results versus the proportion of potentially-detectable isoniazid or fluoroquinolone resistance laboratories are comfortable excluding from the potential benefits of direct LPA DST. For example, for MTBDR*plus* on smear-negatives, C_Tmin_ <29 improves the ratio of actionable-to-non-actionable results by a third and detects >90% of resistance whereas C_Tmin_ <23 permits more than ten actionable results before a non-actionable result occurs yet still detects two-thirds of resistance. For MTBDR*sl*, smaller reductions in ratio compared to MTBDR*plus* occur and more resistance is missed as lower bacillary load specimens are excluded with decreasing C_Tmin_. However, improvements in the ratio for MTBDR*sl* on smear-negatives (3.6-fold or from 1.7 to 5.4, C_Tmin_ <19) would still occur.

Our findings also demonstrate that, where WHO-recommended rapid molecular diagnostic tests are available, smear microscopy, which comes at additional expense and is less accurate at informing when “likely actionable” LPA testing should occur, is increasingly redundant for guiding downstream laboratory decision making given the large range of Xpert C_Tmin_s (and to a lesser extent semi-quantitation categories) within smear-negatives. We therefore suggest PCR test quantitative readouts are used where not all TB-positive specimens undergo automatically reflex DST [this includes MTBDR*plus* for isoniazid resistance, given the prevalence of rifampicin mono-resistant TB (2, 9)].

Our analytical approach can serve as a framework for reflex DSTs other than the LPAs, such as Xpert MTB/XDR (10) and FluoroType MTBDR (11) and others (12), all of which will likely be expensive. Furthermore, the principle of applying molecular (as opposed to visual) quantitative information to determine downstream DST algorithms is agnostic to other frontline TB tests (13, 14) including the Truenat assays (15). Importantly, such frontline tests are increasingly targeting multicopy genes that genotypic DSTs do not include, resulting in large limit of detection differences. Thus, knowing which TB-positive specimens may proceed onward to downstream DST with high actionable result likelihood is a need that will grow.

A strength and limitation is that our study is from a programmatic context, which permitted large sample size, however, the exact thresholds used may require validation in other settings or laboratories. Our study was therefore intended to demonstrate proof-of-concept and illustrate what, purely from a laboratory perspective, such payoffs may look like. Although our findings permit using Xpert to rationally expand the use of existing LPAs to certain paucibacillary specimens ordinarily excluded, we affirm that, resource-permitting, isoniazid and fluroquinolones DST should be attempted directly on any TB-positive rifampicin-resistant specimen irrespective of smear status (16). Hence, our findings will primarily be of interest to settings where direct MTBDR*plus* or MTBDR*sl* testing of smear-negatives is not done (1, 17). Lastly, future work should include Ultra as opposed to Xpert.

In summary, we demonstrated how LPAs may be expanded to a significant proportion of smear-negative patients. Xpert C_Tmin_s or, failing that, Xpert semi-quantitation category is superior to informing reflex LPA testing than smear status, and the utility of molecular quantitative information generated already as part of the TB diagnostic process for informing other reflex tests requires consideration.

## Data Availability

All data produced in the present work are contained in the manuscript.

## Acknowledgements

The authors thank the National Health Laboratory Services, Cape Town, South Africa, and Hain Lifesciences.

## Author Contributions

SP, MdV, GT, and RW conceived experiments. TD and SP provided specimens and data. SP conducted experiments and analysed data. All authors reviewed the manuscript and provided critical input.

